# PARIS Coronary Thrombosis Risk Score Combined With D-dimer to Guide New Oral Anticoagulant Antithrombotic Therapy in Patients With Acute Coronary Syndrome After Percutaneous Coronary Intervention: Rationale and design of the PRIDE-ACS trial

**DOI:** 10.1101/2023.08.01.23293527

**Authors:** Sida Jia, Ying Song, Deshan Yuan, Peizhi Wang, Jingjing Xu, Yan Chen, Ce Zhang, Xueyan Zhao, Jinqing Yuan

## Abstract

**Background:** Residual thrombosis risk is an important contributor to ischemic events in patients with Acute Coronary Syndrome (ACS) after Percutaneous Coronary Intervention (PCI). Although previous studies have shown that rivaroxaban 2.5mg twice daily in ACS patients with high ischemic risk can significantly reduce the risk of ischemic recurrence and mortality, individualized treatment with low-dose rivaroxaban is still rare.

**Aim:** Using D-dimer and PARIS coronary thrombosis risk score to identify ACS patients at high ischemic risk, we aim to investigate whether 3-month low-dose rivaroxaban therapy on the basis of dual antiplatelet therapy (DAPT) could result in reduced ischemic events without increasing bleeding.

**Design:** This study is a multi-center, prospective, open-label, randomized controlled trial involving 3,944 ACS patients undergoing PCI from more than 40 tertiary hospitals in China (ClinicalTrials.gov NCT05638867). Patients with PARIS coronary thrombosis score ≥ 3 and D-dimer ≥ 0.28μg/ml will be 1:1 randomized to experiment group (rivaroxaban 2.5mg twice daily for 3 months on the basis of one-year standard DAPT) or control group (one-year standard DAPT only). The primary endpoint of this study was Major Adverse Cardiovascular and Cerebrovascular Events (MACCE), a composite of death, myocardial infarction, ischemia driven revascularization, stent thrombosis and systemic embolic events. The safety endpoint was BARC type 3 and 5 bleeding events.

**Summary:** In ACS patients with higher PARIS coronary thrombosis risk score and elevated D-dimer level, results of the PRIDE-ACS trial will reveal whether short-duration low-dose rivaroxaban can reduce MACCE events without increasing severe bleeding.

## Background

Despite successful revascularization and effective secondary prevention, rate of recurrent ischemic events remains high for some patients after Percutaneous Coronary Intervention (PCI)(1). Residual thrombotic risk is an important contributor of ischemic events in patients with Acute Coronary Syndrome (ACS), in which thrombin plays an essential role(2). Faced with the trade-off between ischemic and bleeding risk, reducing ischemic events through appropriate anticoagulation has always been a difficult task. Several studies have shown that, albeit with an increased risk of bleeding, the novel oral anticoagulant (NOAC) rivaroxaban can significantly reduce the risk of ischemic recurrence, as well as cardiac and all-cause mortality in patients with coronary heart disease, resulting in net clinical benefit (2–4).

In addition, recent evidences have pointed out that asian population tend to bear higher risk and severity of bleeding than those of other ethnic backgrounds(5–7). Although NOACs seem to have reduced more intracranial hemorrhage risk in the Asian population than in non-Asians compared with warfarin(8), lack of risk assessment tools still limits the clinical usage of NOAC in Chinese ACS patients at high ischemic risk.

Identifying high-risk groups through coronary thrombosis scores and biomarkers enables individualized antithrombotic regimen with NOACs, thereby reducing future ischemic events. D-dimer, a specific degradation product of cross-linked fibrin, is a highly sensitive biomarker for thrombosis, elevated levels of which reflects enhanced coagulation and fibrinolytic activity(9, 10). Our previous study found that baseline D-dimer ≥0.28 μg/ml was independently associated with 2-year all-cause mortality and cardiac death(11). Using the PARIS coronary thrombosis risk score as the basic model, we found that including D-dimer ≥ 0.28ug/ml in the score can significantly improve the predictive power over 2-year cardiac death and coronary thrombosis events (CTEs)(12).

By combining PARIS coronary thrombosis risk score and D-dimer, we can better identify suitable candidates for short-term Triple Antithrombotic Therapy (TAT) with rivaroxaban and DAPT. With the aim to reduce ischemic events without increasing severe bleeding, we designed this multicenter randomized clinical trial (RCT) to evaluate the efficacy and safety of short-term TAT in high-thrombotic-risk ACS population.

## Methods

The PRIDE-ACS trial is an open-label, multi-center, randomized clinical trial. The objective is to evaluate the efficacy and safety of short-term TAT (low-dose rivaroxaban plus DAPT) in comparison with standard DAPT, in ACS patients with PARIS coronary thrombosis score ≥ 3 and baseline D-dimer ≥0.28 μg/ml.

A total of 40 centers are expected to participate in the current study. Inclusion and exclusion criteria are listed in Table 1. Patients are enrolled after a preliminary screening to make sure they meet all inclusion criteria and none of the exclusion criteria.

**Table 1.**
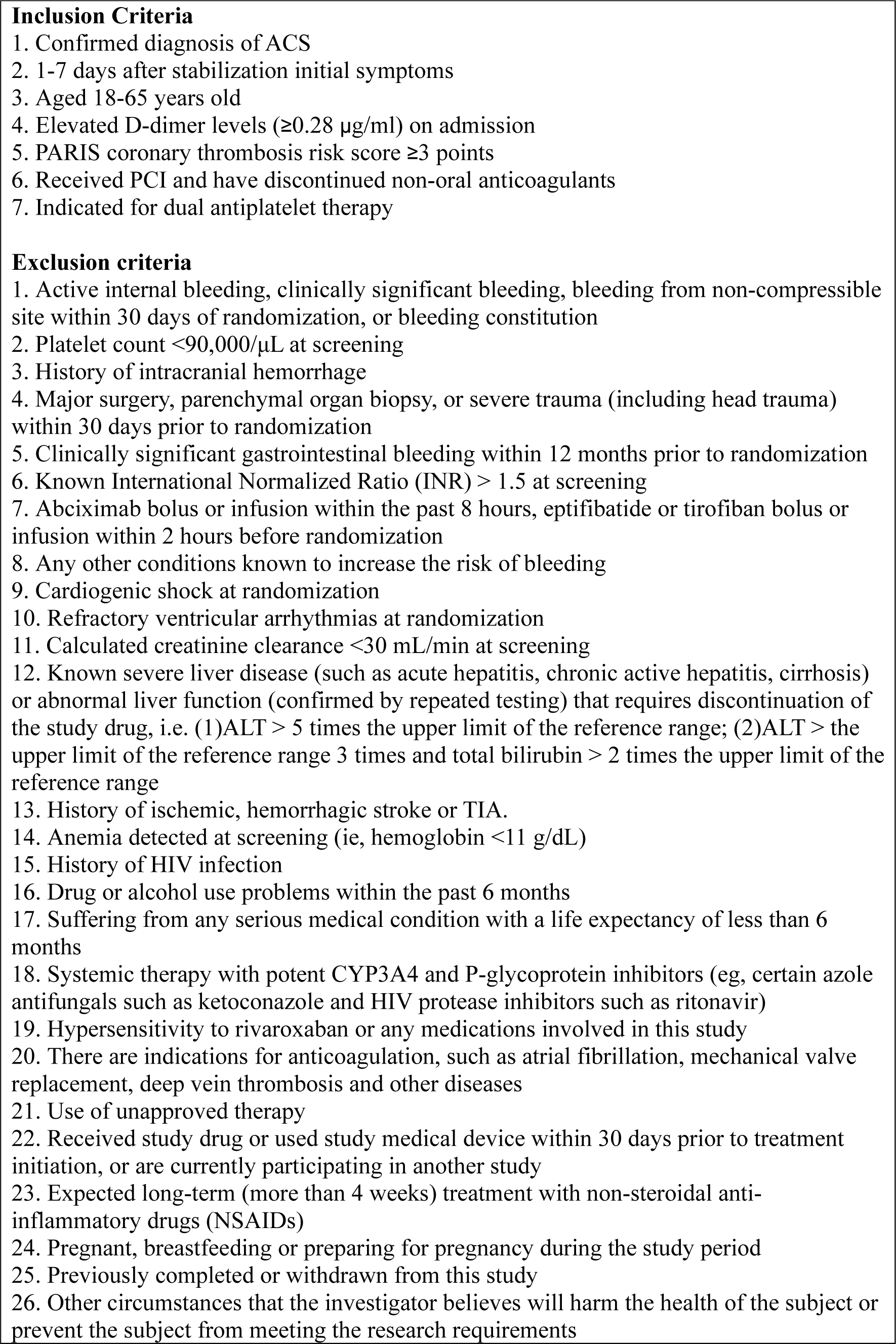
Inclusion and exclusion criteria.

All patients received long-term or loading dose of DAPT before PCI. The regimen is aspirin 75-100 mg once daily for at least 3 days and clopidogrel 75 mg once daily for at least 6 days before PCI. Patients who are not on long-term DAPT were given a loading dose, which contains aspirin 300 mg and clopidogrel 300 mg before PCI, followed by aspirin 75-100 mg and clopidogrel 75 mg orally once a day.

All subjects were screened according to unified inclusion/exclusion criteria. All participants were required to sign an informed consent form before being enrollment. During the randomization process, Interactive Web Respond System (IWRS), a network-based central randomization system, will be used to complete the random allocation of subjects. All patients will be randomized to the experiment group or the control group at 1:1 ratio. Patients assigned to the experiment group will receive TAT for the first 3 months (aspirin 75-100 mg QD, clopidogrel 75 mg QD and rivaroxaban 2.5 mg BID), followed by conventional DAPT for 9 months. Patients assigned to the control group will receive conventional DAPT for 1 year. During follow-up, the basic DAPT regimen of the two groups remained unchanged. D-dimer level will be re-examined at 3, 6, and 12 months (Figure 1).

Baseline physical examination results, medical history, ECGs, laboratory testing results, as well as concomitant medications are collected on admission according to Table 2. Clinic follow-up visits are scheduled at 3 months (±1 week), 6 months (±1 month), and 12 months (±1 month). Coagulation function test, complete blood count, urinalysis, stool analysis, biochemical test and biomarkers test will be re-checked at 3-month and 6-month follow-up. Evaluation of compliance, concomitant medication, and documentation of adverse events will be performed during each follow-up.

**Table 2.**
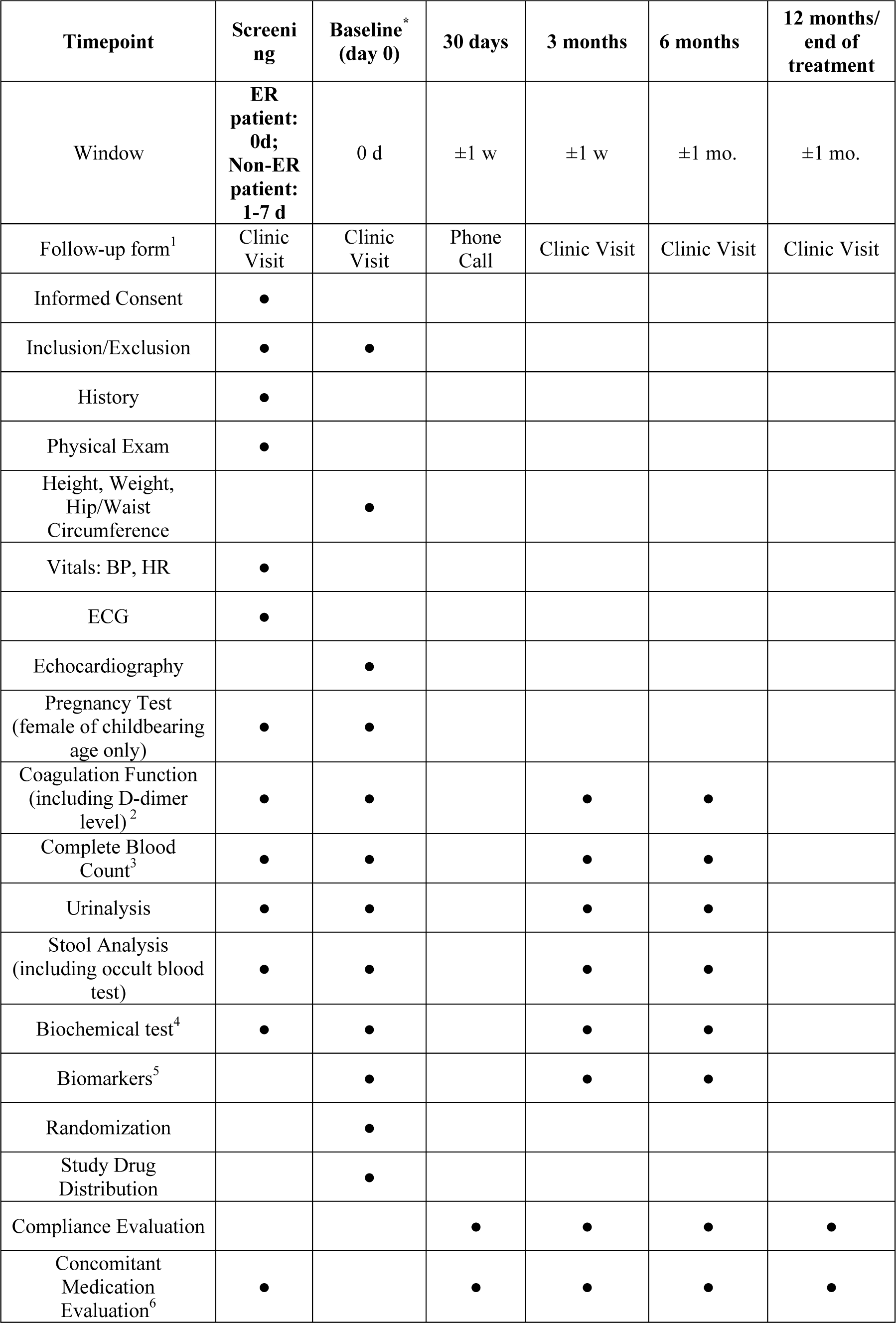
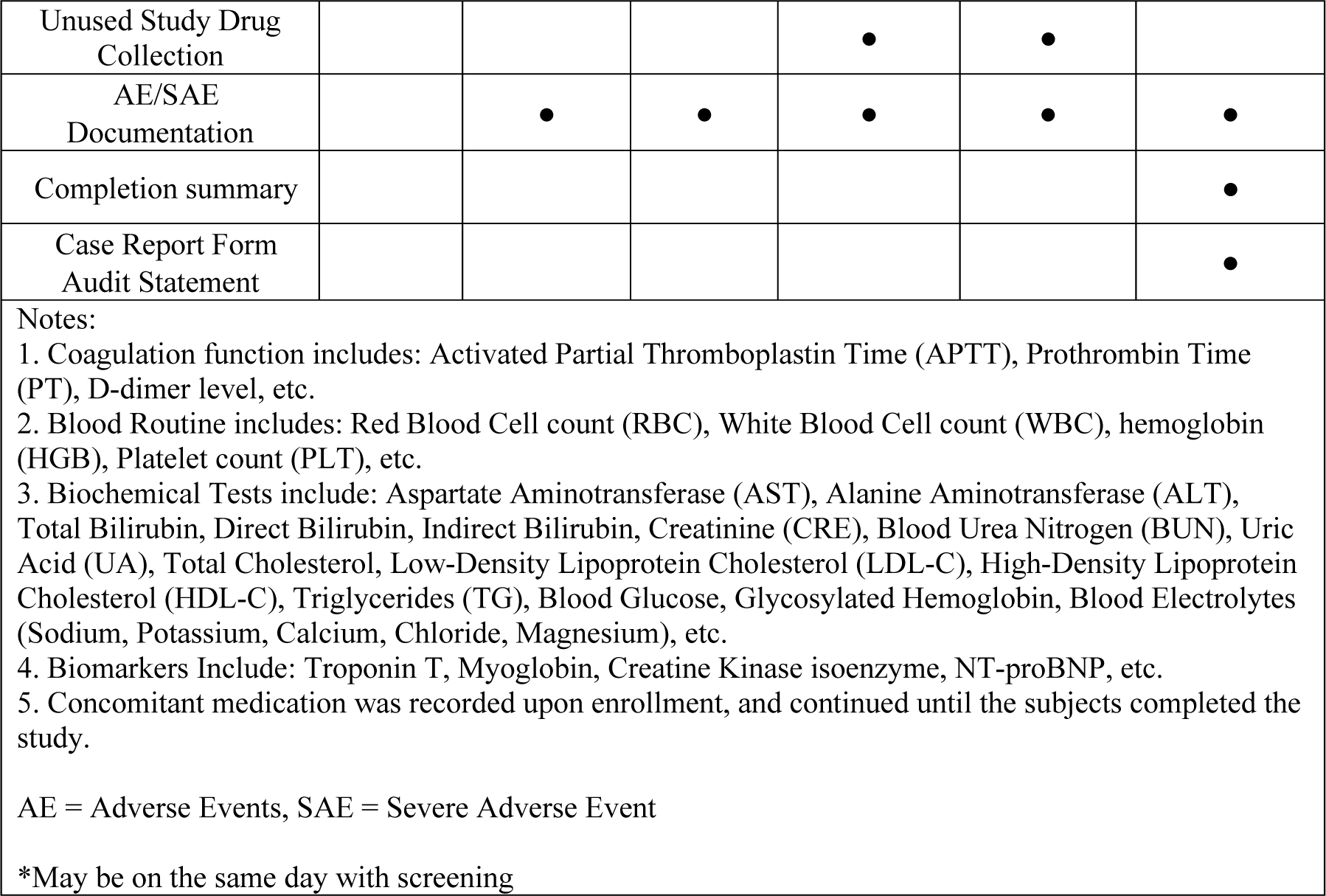
Schedule of data acquisition.

| Timepoint | Screening | Baseline*<br>(day 0) | 30 days | 3 months | 6 months | 12 months/<br>end of treatment |
| --- | --- | --- | --- | --- | --- | --- |
| Window | **ER patient:**<br>**0d;**<br>**Non-ER patient:**<br>**1-7 d** | 0 d | ±1 w | ±1 w | ±1 mo. | ±1 mo. |
| Follow-up form<sup>1</sup> | Clinic Visit | Clinic Visit | Phone Call | Clinic Visit | Clinic Visit | Clinic Visit |
| Informed Consent | • |  |  |  |  |  |
| Inclusion/Exclusion | • | • |  |  |  |  |
| History | • |  |  |  |  |  |
| Physical Exam | • |  |  |  |  |  |
| Height, Weight, Hip/Waist Circumference |  | • |  |  |  |  |
| Vitals: BP, HR | • |  |  |  |  |  |
| ECG | • |  |  |  |  |  |
| Echocardiography |  | • |  |  |  |  |
| Pregnancy Test (female of childbearing age only) | • | • |  |  |  |  |
| Coagulation Function (including D-dimer level)<sup>2</sup> | • | • |  | • | • |  |
| Complete Blood Count<sup>3</sup> | • | • |  | • | • |  |
| Urinalysis | • | • |  | • | • |  |
| Stool Analysis (including occult blood test) | • | • |  | • | • |  |
| Biochemical test<sup>4</sup> | • | • |  | • | • |  |
| Biomarkers<sup>5</sup> |  | • |  | • | • |  |
| Randomization |  | • |  |  |  |  |
| Study Drug Distribution |  | • |  |  |  |  |
| Compliance Evaluation |  |  | • | • | • | • |
| Concomitant Medication Evaluation<sup>6</sup> | • |  | • | • | • | • |
| Unused Study Drug Collection |  |  |  | • | • |  |
| AE/SAE Documentation |  | • | • | • | • | • |
| Completion summary |  |  |  |  |  | • |
| Case Report Form Audit Statement |  |  |  |  |  | • |

Notes:
1. Coagulation function includes: Activated Partial Thromboplastin Time (APTT), Prothrombin Time (PT), D-dimer level, etc. 2. Blood Routine includes: Red Blood Cell count (RBC), White Blood Cell count (WBC), hemoglobin (HGB), Platelet count (PLT), etc. 3. Biochemical Tests include: Aspartate Aminotransferase (AST), Alanine Aminotransferase (ALT), Total Bilirubin, Direct Bilirubin, Indirect Bilirubin, Creatinine (CRE), Blood Urea Nitrogen (BUN), Uric Acid (UA), Total Cholesterol, Low-Density Lipoprotein Cholesterol (LDL-C), High-Density Lipoprotein Cholesterol (HDL-C), Triglycerides (TG), Blood Glucose, Glycosylated Hemoglobin, Blood Electrolytes (Sodium, Potassium, Calcium, Chloride, Magnesium), etc. 4. Biomarkers Include: Troponin T, Myoglobin, Creatine Kinase isoenzyme, NT-proBNP, etc. 5. Concomitant medication was recorded upon enrollment, and continued until the subjects completed the study.
AE = Adverse Events, SAE = Severe Adverse Event
\*May be on the same day with screening

In terms of concomitant medication, the guideline-recommended medications for secondary prevention remain unchanged for all patients during follow-up. Anticoagulants contraindicated in this trial include: (1) other oral anticoagulants such as warfarin, dabigatran, apixaban, etc.; (2) other intravenous anticoagulants such as heparin, low molecular weight heparin, bivalirudin, fondaparinux sodium, etc. Furthermore, strong inducers or inhibitors of P-gp and/or CYP3A4 cannot be used in combination with rivaroxaban according to guideline recommendations(13). Patients who were already on the medications listed above will be excluded. If patients enrolled are clinically indicated to use the medications listed above, the trial rivaroxaban will be discontinued after discussion by the expert committee. Investigators and follow-up staff will keep detailed records of discontinuation of the trial rivaroxaban.

The follow-up period of this study is 12 months. The primary endpoint is Major Adverse Cardiovascular and Cerebrovascular Events (MACCE), a composite of all-cause death, myocardial infarction, stroke, ischemia-driven revascularization, stent thrombosis and systemic embolism composite events. Secondary endpoints include all-cause death, cardiac death, myocardial infarction, stroke, ischemia-driven revascularization, stent thrombosis, systemic embolism and net clinical adverse events (NACE), a composite of MACCE or BARC 3, 5 bleeding events). The safety endpoint is BARC 3, 5 bleeding events. Detailed definition of the endpoints are listed in the Supplementary Table 1.

In this study, Adverse Events (AEs) refer to all adverse medical events that occur after subjects receive trial rivaroxaban, which can be manifested as symptoms, signs, diseases or abnormal laboratory tests, but are not necessarily causally related to trial rivaroxaban. All AEs will be documented in detail, including but not limited to name, date, severity and whether it is related to trial rivaroxaban. All patients with AEs will be followed-up until recovery or stabilization. AEs that meet one or more of the following criteria is defined as a severe adverse event (SAE): (1) death, (2) life-threatening situation, (3) permanent or severe disabilities, (4) rehospitalization or prolonged hospital stay, (5) congenital anomalies or birth defects. Any SAEs will be reported to the investigators team, the sponsor, the ethics committee, and related administrative departments within 24 hours.

Suspected and Unexpected Serious Adverse Reactions (SUSAR) refer to adverse reactions whose nature and severity exceed the existing data and information, such as the Investigator’s Manual or the Summary of Product Characteristics. After the research team is informed of an SAE, a comprehensive analysis and evaluation of the SAE should be carried out immediately to determine whether the SAE is a SUSAR and therefore should be reported to the ethics committee.

The sample size of this study was calculated using SAS 9.4 software. With reference to previous studies, we expect the incidence of MACCE events at 12 months to be 13% and 10% in the control group(2, 14) and experiment group(2, 14, 15), respectively. Based on this assumption and the superiority study design, an estimated sample size of 1,774 in each group would give 80% (2-sided α = 0.05 and β = 0.20) statistical power to detect a significant difference. Considering a dropout rate of 10%, the total target sample size is set at 3,944 patients (1,972 in each arm).

The primary analysis will be performed according to a modified intention-to-treat principle. The modified intention-to-treat population consists of patients undergoing randomization but excluding those with randomization error or informed consent withdrawal prior to baseline clinic visit. Primary and major secondary end points will also be analyzed in both per-protocol and as-treated populations. The per-protocol population consists of patients who were successfully randomized and underwent the assigned treatment excluding those with major protocol deviations. The as treated population consists of patients who are analyzed according to the actual strategy used for treatment rather than their randomization assignment.

For patient baseline data, categorical variables will be presented as frequencies and percentages. Continuous variables will be displayed as mean ± standard deviation or median (interquartile range), as appropriate. Continuous variables will be analyzed for normality using the Kolmogorov-Smirnov test. Chi-square test or Fisher’s exact test will be used in comparisons of categorical variables between groups, whereas the Student t-test or Wilcoxon rank-sum test will be used in comparison of continuous variables, as appropriate. First-time event rates were estimated for each group using the Kaplan-Meier method and compared by the log-rank test. Differences between groups are estimated by hazard ratios (HRs) with 95% confidence intervals using Cox proportional hazards models. A two-tailed p-value <0.05 is considered statistically significant. Statistical analysis will be performed using SPSS version 24 software (or higher version; SPSS Inc.).

The Steering Committee of the present study, chaired by Jinqing Yuan, MD, FACC, FESC, is responsible for devising study protocol, overseeing the operation of this trial, as well as drafting and preparation of potential manuscripts and other publications from this study.

Clinical Events Committee (CEC) will be established to make independent judgments on clinical endpoints. The CEC consists of independent cardiovascular experts and was blinded to patient grouping information. The CEC is responsible for determining whether a reported clinical event constitutes an endpoint event, according to the classification criteria specified in Supplementary Table 2. Data related to clinical endpoints were uniformly collected and organized by follow-up staff blinded to grouping as required.

Data Safety Monitoring Board (DSMB) is responsible for regularly reviewing overall and individual patient data related to safety, data integrity and overall operation of the study to ensure subject safety. The DSMB consists of at least five members, including at least two cardiologists, one neurologist, one statistician, and another independent statistician who is responsible for the statistics required for DSMB meetings, but will not participate in decision-making votes. The DSMB’s detailed charter will be drawn up at its first meeting. After each meeting, the DSMB evaluates the trial benefit/risk ratio, reaches consensus through discussion, and reports to the investigator its recommendations to continue the trial with or without modification, or terminate the trial.

Institutional review board approval by the ethics committee of Fuwai Hospital was obtained on April 22, 2023 (Approval No. 2023-1980). The approved study protocol, informed consent and other relevant materials are sent to other participating centers. Before patient enrollment and randomization, all centers should gain approval from their own ethics committee. The study protocol was registered on ClinicalTrials.gov (no. NCT05638867) in November 2022.

## Discussion

Even after successful revascularization and effective secondary prevention, recurrent ischemic events still occur in 5% to 10% of patients after PCI annually(1). In patients with triple-vessel Coronary Artery Disease (CAD), our previous cohort study found that despite revascularization and/or optimal medical therapy according to guidelines, over a median follow-up of over 7 years, about a quarter of patients still suffered from MACCEs (16), with even higher rate of MACCEs in patients presenting as non-ST elevation ACS (NSTE-ACS)(17). Another cohort of ours including 10,724 consecutive patients undergoing PCI showed that 21.6% of patients had MACCE during 5-year follow-up(18). Since residual thrombosis risk stands out among various causes of ischemic events after PCI, reducing residual thrombosis risk is crucial in improving patient prognosis.

Both platelet and thrombin play a central role in the process of thrombus formation. Previous study showed that thrombin activity continues to rise within 1 year after myocardial infarction and within 6 months after the onset of unstable angina(2). How to properly inhibit thrombin to reduce ischemic events in ACS patients has been under heated debate.

Rivaroxaban, a selective factor Xa inhibitor, inhibits thrombin formation by blocking both intrinsic and extrinsic coagulation cascades. The ATLAS ACS 2-TIMI 51 study(4) found that in patients with ACS, rivaroxaban 2.5 mg twice daily brings a clear net clinical benefit by significantly reducing the risk of ischemic recurrence, cardiac death and all-cause death. Although the rivaroxaban group had a significantly higher risk of bleeding than the control group (1.8% vs. 0.6%) possibly due to long duration of anticoagulation (13.1 months on average), no increase in fatal bleeding was observed. In patients with stable CAD or peripheral artery disease, the COMPASS study(4) found that Dual Path Inhibition (DPI) with rivaroxaban and aspirin reduced adverse cardiovascular events compared with aspirin alone. In terms of safety, despite increased International Society on Thrombosis and Haemostasis (ISTH) major bleeding in DPI group, fatality and major organ bleeding was comparable with aspirin alone, leading to greater net clinical benefit in high ischemic-risk patients(4). A subgroup analysis of patients with stable CAD and PCI history revealed similar results (19). Recent European guidelines on STEMI(20) and NSTE-ACS(21) both recommended that low-dose rivaroxaban (2.5 mg twice daily) can be considered in combination with aspirin and clopidogrel for patients at low bleeding risk (IIb B). In ACS setting, using low-dose rivaroxaban is still debatable and individualized rivaroxaban use is rare. Future randomized clinical trials are warranted to help identify suitable ACS population for precise anticoagulation.

Choosing the correct anticoagulant, dose and duration of treatment is crucial to reducing the risk of thrombosis and bleeding. The ATLAS ACS2-TIMI51 study confirmed that TAT (dual antiplatelet therapy combined with rivaroxaban) can significantly reduce ischemic risk, but increases the risk of non-CABG-related TIMI major bleeding (HR: 3.46, 95% CI: 2.08–5.77, p<0.001)(2). On primary efficacy endpoint and cardiovascular death, the cumulative incidence curves of rivaroxaban and placebo group began to separate at 30 days during follow-up, after which the curves seemed parallel. In terms of safety endpoints, compared with the triple antithrombotic group, the risk of major bleeding at 3 months was significantly lower than that at 6 months in the DAPT group (0.5% vs 0.8%)(2). In other words, long-term TAT seems ineffective in decreasing thrombotic events, but contrarily might bring additional bleeding risk. Therefore, in a population with high ischemic risk, shorter-duration TAT might be promising in improving net clinical outcomes. Based on the results from ATLAS ACS2-TIMI51 trial(2), the ATLAS ACS-TIMI46 trial(3) and analysis from our own cohort of Chinese PCI patients(22), we have limited candidates for TAT to high-thrombotic-risk ACS patients, and shortened TAT duration to 3 months in PRIDE-ACS trial.

D-dimer is a specific degradation product of cross-linked fibrin and a highly sensitive biomarker of thrombus formation(9, 23). In recent years, multiple studies have shown that D-dimer levels are associated with long-term adverse cardiovascular events in patients with ACS and stable CAD. In patients with stable CAD, the LIPID study found that elevated D-dimer levels not only increased major coronary events, MACE and deep vein thrombosis, but was also an independent risk factor for adverse events including all-cause death and cardiac death(24). A subgroup analysis of ATLAS ACS-TIMI 46 trial showed that in ACS patients, elevated baseline D-dimer levels were associated with an increased risk of composite adverse events within 6 months, and that rivaroxaban was associated with decreased D-dimer levels(25). Consistently, our cohort study in patients undergoing PCI found that baseline D-dimer levels ≥ 0.28μg/ml were independently associated with 2-year all-cause and cardiac death(26). Nevertheless, due to low specificity, D-dimer alone has limited power in risk stratification and prognosis assessment in ACS patients.

Compared with a single biomarker, scoring models can better identify high-risk patients and predict short-term and long-term adverse events. However, there still lacks ideal thrombosis risk prediction model in clinical practice. Currently, the PARIS coronary thrombosis risk score recommended by the guidelines is a common scoring tool used to predict the risk of ischemic adverse events in patients taking dual antiplatelet therapy after PCI(27). The risk score was derived from the PARIS registry, with one of the primary study endpoints being coronary thrombotic events (CTEs, a composite endpoint of stent thrombosis and myocardial infarction). The validation cohort revealed a modest predictive ability for the risk of thrombosis (C statistic = 0.65)(27). In Chinese patients undergoing PCI, validation in our cohort also revealed limited predictive power of PARIS thrombosis risk score over CTEs(28) and death(11), with the c-statistic being 0.621 and 0.607 respectively. One of the limitations of the PARIS coronary thrombosis score is that no biomarkers are incorporated into the scoring system to predict thrombotic events.

Preliminary results of our cohort study showed that in patients with PARIS coronary thrombosis score ≥ 3 points, adding D-dimer ≥ 0.28ug/ml can improve the predictive value of PARIS coronary thrombosis score for 2-year cardiac death and CTEs (c-statistics increased to 0.721 and 0.655, both p<0.05), with a significant improvement in net reclassification index (p<0.05)(12). Furthermore, in patients with PARIS coronary thrombosis score ≥ 3 points, patients with D-dimer ≥ 0.28ug/ml had a low and similar rate of BARC type 3 and 5 bleeding compared with the D-dimer <0.28ug/ml counterparts (0.6% vs. 0.5%, p>0.05)(12). It seems combining D-dimer and PARIS coronary thrombosis score, a thrombosis biomarker with a thrombosis risk score, can better identify ACS patients at high thrombosis risk, the ideal population for precise short-term TAT with rivaroxaban.

In conclusion, by combining PARIS coronary thrombosis risk score and D-dimer, the current study aim to identify ACS patients at high thrombosis risk after PCI, and investigate whether 3-month TAT with low-dose rivaroxaban in this specific population could reduce ischemia without increasing severe bleeding. The results of this study are expected to provide strong evidence on rivaroxaban therapy in selected high-risk ACS patients after PCI, and will hopefully help improve clinical outcomes in this specific population.

## Funding

The present study is supported by the National Clinical Research Center for Cardiovascular Diseases, Fuwai Hospital, Chinese Academy of Medical Sciences (Grant No.

NCRC2022003).

## Data Availability Statements

No new data were generated or analysed in support of this research.

## Reference

1. Madhavan MV, Kirtane AJ, Redfors B, Genereux P, Ben-Yehuda O, Palmerini T, Benedetto U, Biondi-Zoccai G, Smits PC, von Birgelen C, Mehran R, McAndrew T, Serruys PW, Leon MB, Pocock SJ, Stone GW. Stent-Related Adverse Events >1 Year After Percutaneous Coronary Intervention. J Am Coll Cardiol 2020; 75(6):590–604.

2. Mega JL, Braunwald E, Wiviott SD, Bassand JP, Bhatt DL, Bode C, Burton P, Cohen M, Cook-Bruns N, Fox KA, Goto S, Murphy SA, Plotnikov AN, Schneider D, Sun X, Verheugt FW, Gibson CM, Investigators AAT. Rivaroxaban in patients with a recent acute coronary syndrome. N Engl J Med 2012; 366(1):9–19.

3. Mega JL, Braunwald E, Mohanavelu S, Burton P, Poulter R, Misselwitz F, Hricak V, Barnathan ES, Bordes P, Witkowski A, Markov V, Oppenheimer L, Gibson CM, group AA-Ts. Rivaroxaban versus placebo in patients with acute coronary syndromes (ATLAS ACS-TIMI 46): a randomised, double-blind, phase II trial. Lancet 2009; 374(9683):29–38.

4. Eikelboom JW, Connolly SJ, Bosch J, Dagenais GR, Hart RG, Shestakovska O, Diaz R, Alings M, Lonn EM, Anand SS, Widimsky P, Hori M, Avezum A, Piegas LS, Branch KRH, Probstfield J, Bhatt DL, Zhu J, Liang Y, Maggioni AP, Lopez-Jaramillo P, O’Donnell M, Kakkar AK, Fox KAA, Parkhomenko AN, Ertl G, Störk S, Keltai M, Ryden L, Pogosova N, Dans AL, Lanas F, Commerford PJ, Torp-Pedersen C, Guzik TJ, Verhamme PB, Vinereanu D, Kim J-H, Tonkin AM, Lewis BS, Felix C, Yusoff K, Steg PG, Metsarinne KP, Cook Bruns N, Misselwitz F, Chen E, Leong D, Yusuf S. Rivaroxaban with or without Aspirin in Stable Cardiovascular Disease. The New England Journal of Medicine 2017; 377(14):1319–1330.

5. Lee SR, Choi EK, Kwon S, Jung JH, Han KD, Cha MJ, Oh S, Lip GYH. Oral Anticoagulation in Asian Patients With Atrial Fibrillation and a History of Intracranial Hemorrhage. Stroke 2020; 51(2):416–423.

6. Bako AT, Pan AP, Potter T, Meeks JR, Cainzos-Achirica M, Woo D, Vahidy FS. Demographic Characteristics and Clinical Outcomes of Asian American and Pacific Islander Patients With Primary Intracerebral Hemorrhage. JAMA Netw Open 2021; 4(12):e2138786.

7. Tse WC, Grey C, Harwood M, Jackson R, Kerr A, Mehta S, Poppe K, Pylypchuk R, Wells S, Selak V. Risk of major bleeding by ethnicity and socioeconomic deprivation among 488,107 people in primary care: a cohort study. BMC Cardiovasc Disord 2021; 21(1):206.

8. Wang KL, Lip GY, Lin SJ, Chiang CE. Non-Vitamin K Antagonist Oral Anticoagulants for Stroke Prevention in Asian Patients With Nonvalvular Atrial Fibrillation: Meta-Analysis. Stroke 2015; 46(9):2555–61.

9. Johnson ED, Schell JC, Rodgers GM. The D-dimer assay. American Journal of Hematology 2019; 94(7):833–839.

10. Ariens RA, de Lange M, Snieder H, Boothby M, Spector TD, Grant PJ. Activation markers of coagulation and fibrinolysis in twins: heritability of the prethrombotic state. Lancet 2002; 359(9307):667–71.

11. Zhao X, Li J, Tang X, Xian Y, Jiang L, Chen J, Gao L, Gao Z, Qiao S, Yang Y, Gao R, Xu B, Yuan J. Prognostic Value of the PARIS Thrombotic Risk Score for 2-Year Mortality After Percutaneous Coronary Intervention. Clin Appl Thromb Hemost 2019; 25:1076029619853638.

12. Jia S, Deshan Y, Song Y, Xu J, Zhao X, Yuan J. INDEPENDENT AND INCREMENTAL PROGNOSTIC VALUE OF D-DIMER IN ACUTE CORONARY SYNDROME PATIENTS UNDERGOING PCI: DATA FROM A LARGE PROSPECTIVE COHORT STUDY. Journal of the American College of Cardiology 2023; 81(8_Supplement):1184–1184.

13. Steffel J, Collins R, Antz M, Cornu P, Desteghe L, Haeusler KG, Oldgren J, Reinecke H, Roldan-Schilling V, Rowell N, Sinnaeve P, Vanassche T, Potpara T, Camm AJ, Heidbüchel H. 2021 European Heart Rhythm Association Practical Guide on the Use of Non-Vitamin K Antagonist Oral Anticoagulants in Patients with Atrial Fibrillation. Europace : European Pacing, Arrhythmias, and Cardiac Electrophysiology : Journal of the Working Groups On Cardiac Pacing, Arrhythmias, and Cardiac Cellular Electrophysiology of the European Society of Cardiology 2021; 23(10):1612–1676.

14. Tang Y-D, Wang W, Yang M, Zhang K, Chen J, Qiao S, Yan H, Wu Y, Huang X, Xu B, Gao R, Yang Y. Randomized Comparisons of Double-Dose Clopidogrel or Adjunctive Cilostazol Versus Standard Dual Antiplatelet in Patients With High Posttreatment Platelet Reactivity: Results of the CREATIVE Trial. Circulation 2018; 137(21):2231–2245.

15. Gibson CM, Mehran R, Bode C, Halperin J, Verheugt FW, Wildgoose P, Birmingham M, Ianus J, Burton P, van Eickels M, Korjian S, Daaboul Y, Lip GYH, Cohen M, Husted S, Peterson ED, Fox KA. Prevention of Bleeding in Patients with Atrial Fibrillation Undergoing PCI. The New England Journal of Medicine 2016; 375(25):2423–2434.

16. Zhang C, Jiang L, Xu L, Tian J, Liu J, Zhao X, Feng X, Wang D, Zhang Y, Sun K, Xu B, Zhao W, Hui R, Gao R, Yuan J, Song L. Implications of N-terminal pro-B-type natriuretic peptide in patients with three-vessel disease. European Heart Journal 2019; 40(41):3397–3405.

17. Jia S, Zhang C, Jiang L, Xu L, Tian J, Zhao X, Feng X, Wang D, Zhang Y, Sun K, Xu J, Liu R, Xu B, Zhao W, Hui R, Gao R, Gao Z, Yuan J, Song L. Comparison of Percutaneous Coronary Intervention, Coronary Artery Bypass Grafting and Medical Therapy in Non-ST Elevation Acute Coronary Syndrome Patients With 3-Vessel Disease. Circulation Journal : Official Journal of the Japanese Circulation Society 2020; 84(10):1718–1727.

18. Jia S, Li J, Zhang C, Liu Y, Yuan D, Xu N, Zhao X, Gao R, Yang Y, Xu B, Gao Z, Yuan J, Zhang Y. Long-Term Prognosis of Moderate to Severe Coronary Artery Calcification in Patients Undergoing Percutaneous Coronary Intervention. Circulation Journal : Official Journal of the Japanese Circulation Society 2020; 85(1):50–58.

19. Bainey KR, Welsh RC, Connolly SJ, Marsden T, Bosch J, Fox KAA, Steg PG, Vinereanu D, Connolly DL, Berkowitz SD, Foody JM, Probstfield JL, Branch KR, Lewis BS, Diaz R, Muehlhofer E, Widimsky P, Yusuf S, Eikelboom JW, Bhatt DL. Rivaroxaban Plus Aspirin Versus Aspirin Alone in Patients With Prior Percutaneous Coronary Intervention (COMPASS-PCI). Circulation 2020; 141(14):1141–1151.

20. Ibanez B, James S, Agewall S, Antunes MJ, Bucciarelli-Ducci C, Bueno H, Caforio ALP, Crea F, Goudevenos JA, Halvorsen S, Hindricks G, Kastrati A, Lenzen MJ, Prescott E, Roffi M, Valgimigli M, Varenhorst C, Vranckx P, Widimský P, Group ESD. 2017 ESC Guidelines for the management of acute myocardial infarction in patients presenting with ST-segment elevation: The Task Force for the management of acute myocardial infarction in patients presenting with ST-segment elevation of the European Society of Cardiology (ESC). European Heart Journal 2017; 39(2):119–177.

21. Collet J-P, Thiele H, Barbato E, Barthélémy O, Bauersachs J, Bhatt DL, Dendale P, Dorobantu M, Edvardsen T, Folliguet T, Gale CP, Gilard M, Jobs A, Jüni P, Lambrinou E, Lewis BS, Mehilli J, Meliga E, Merkely B, Mueller C, Roffi M, Rutten FH, Sibbing D, Siontis GCM, Chettibi M, Hayrapetyan HG, Metzler B, Najafov R, Stelmashok VI, Claeys M, Kušljugić Z, Gatzov PM, Skoric B, Panayi G, Mates M, Sorensen R, Shokry K, Marandi T, Kajander OA, Commeau P, Aladashvili A, Massberg S, Nikas D, Becker D, Guðmundsdóttir IJ, Peace AJ, Beigel R, Indolfi C, Aidargaliyeva N, Elezi S, Beishenkulov M, Maca A, Gustiene O, Degrell P, Cassar Maempel A, Ivanov V, Damman P, Kedev S, Steigen TK, Legutko J, Morais J, Vinereanu D, Duplyakov D, Zavatta M, Pavlović M, Orban M, Bunc M, Ibañez B, Hofmann R, Gaemperli O, Marjeh YB, Addad F, Tutar E, Parkhomenko A, Karia N, Group ESD. 2020 ESC Guidelines for the management of acute coronary syndromes in patients presenting without persistent ST-segment elevation: The Task Force for the management of acute coronary syndromes in patients presenting without persistent ST-segment elevation of the European Society of Cardiology (ESC). European Heart Journal 2020; 42(14):1289–1367.

22. Sida Jia YD, Ying Song, Jingjing Xu, Xueyan Zhao, Jinqing Yuan. INDEPENDENT AND INCREMENTAL PROGNOSTIC VALUE OF D-DIMER IN ACUTE CORONARY SYNDROME PATIENTS UNDERGOING PCI: DATA FROM A LARGE PROSPECTIVE COHORT STUDY. In; 2023.

23. Ariëns RAS, de Lange M, Snieder H, Boothby M, Spector TD, Grant PJ. Activation markers of coagulation and fibrinolysis in twins: heritability of the prethrombotic state. Lancet (London, England) 2002; 359(9307):667–671.

24. Simes J, Robledo KP, White HD, Espinoza D, Stewart RA, Sullivan DR, Zeller T, Hague W, Nestel PJ, Glasziou PP, Keech AC, Elliott J, Blankenberg S, Tonkin AM. D-Dimer Predicts Long-Term Cause-Specific Mortality, Cardiovascular Events, and Cancer in Patients With Stable Coronary Heart Disease: LIPID Study. Circulation 2018; 138(7):712–723.

25. AlKhalfan F, Kerneis M, Nafee T, Yee MK, Chi G, Plotnikov A, Braunwald E, Gibson CM. D-Dimer Levels and Effect of Rivaroxaban on Those Levels and Outcomes in Patients With Acute Coronary Syndrome (An ATLAS ACS-TIMI 46 Trial Substudy). The American Journal of Cardiology 2018; 122(9):1459–1464.

26. Zhao X, Li J, Tang X, Jiang L, Chen J, Qiao S, Yang Y, Gao R, Xu B, Yuan J. D-dimer as a thrombus biomarker for predicting 2-year mortality after percutaneous coronary intervention. Therapeutic Advances In Chronic Disease 2020; 11:2040622320904302.

27. Baber U, Mehran R, Giustino G, Cohen DJ, Henry TD, Sartori S, Ariti C, Litherland C, Dangas G, Gibson CM, Krucoff MW, Moliterno DJ, Kirtane AJ, Stone GW, Colombo A, Chieffo A, Kini AS, Witzenbichler B, Weisz G, Steg PG, Pocock S. Coronary Thrombosis and Major Bleeding After PCI With Drug-Eluting Stents: Risk Scores From PARIS. Journal of the American College of Cardiology 2016; 67(19):2224–2234.

28. Zhao X-Y, Li J-X, Tang X-F, Xu J-J, Song Y, Jiang L, Chen J, Song L, Gao L-J, Gao Z, Qiao S-B, Yang Y-J, Gao R-L, Xu B, Yuan J-Q. Validation of Predictive Value of Patterns of Nonadherence to Antiplatelet Regimen in Stented Patients Thrombotic Risk Score in Chinese Population Undergoing Percutaneous Coronary Intervention: A Prospective Observational Study. Chinese Medical Journal 2018; 131(22):2699–2704.

